# Cross-reactive antibodies after SARS-CoV-2 infection and vaccination

**DOI:** 10.1101/2021.05.26.21256092

**Authors:** Marloes Grobben, Karlijn van der Straten, Philip J.M. Brouwer, Mitch Brinkkemper, Pauline Maisonnasse, Nathalie Dereuddre-Bosquet, Judith A. Burger, Meliawati Poniman, Melissa Oomen, Dirk Eggink, Tom P.L. Bijl, Hugo D.G. van Willigen, Elke Wynberg, Bas J. Verkaik, Orlane J.A. Figaroa, Peter J. de Vries, Tessel M. Boertien, Roger Le Grand, Menno D. de Jong, Maria Prins, Amy W. Chung, Godelieve J. de Bree, Rogier W. Sanders, Marit J. van Gils

**Author notes:** Corresponding author: (M.J.v.G). These authors contributed equally (Marloes Grobben, Karlijn van der Straten).

## Abstract

Current SARS-CoV-2 vaccines are losing efficacy against emerging variants and may not protect against future novel coronavirus outbreaks, emphasizing the need for more broadly protective vaccines. To inform the development of a pan-coronavirus vaccine, we investigated the presence and specificity of cross-reactive antibodies against the spike (S) proteins of human coronaviruses (hCoV) after SARS-CoV-2 infection and vaccination. We found an 11 to 123-fold increase in antibodies binding to SARS-CoV and MERS-CoV as well as a 2 to 4-fold difference in antibodies binding to seasonal hCoVs in COVID-19 convalescent sera compared to pre-pandemic healthy donors, with the S2 subdomain of the S protein being the main target for cross-reactivity. In addition, we detected cross-reactive antibodies to all hCoV S proteins after SARS-CoV-2 S protein immunization in macaques, with higher responses for hCoV more closely related to SARS-CoV-2. These findings support the feasibility of and provide guidance for development of a pan-coronavirus vaccine.

## Introduction

One and a half year after the emergence of Severe Acute Respiratory Coronavirus 2 (SARS-CoV-2), the causative agent of coronavirus infectious disease 2019 (COVID-19), the pandemic is still a major global issue with unprecedented consequences on healthcare systems and economies. Following the rapid initiation of many COVID-19 vaccine studies, the first vaccines are already FDA/EMA approved and mass vaccination campaigns are rolled out to hopefully soon subdue this pandemic^1–4^. However, these vaccines might have reduced efficacy against emerging SARS-CoV-2 variants, as was already shown for several approved vaccines when tested for safety and effectiveness against the B1.351 (501Y.V2) SARS-CoV-2 variant which is currently dominating the epidemic in South Africa^5,6,7^. To anticipate emerging variants, more broadly protective SARS-CoV-2 vaccines are desirable. However, the ultimate goal should be a pan-coronavirus vaccine, which would be able to induce broad immunity and protection against multiple coronaviruses, thereby preparing us for future outbreaks.

New coronaviruses that infect humans (hCoVs) emerge frequently and SARS-CoV-2 is the fifth to be discovered in less than two decades, bringing the total up to seven^8^. These include the seasonal betacoronaviruses hCoV-OC43 and hCoV-HKU1 as well as alphacoronaviruses hCoV-NL63 and hCoV-229E, which usually cause mild respiratory symptoms and account for approximately 5-30% of common colds during the winter season^9,10^. In contrast, Severe Acute Respiratory Syndrome (SARS)-CoV and Middle Eastern Respiratory Syndrome (MERS)-CoV are far more pathogenic and led to epidemics in 2003 and 2013-2015, respectively^11,12^. The currently circulating SARS-CoV-2 is less pathogenic compared to SARS-CoV and MERS-CoV, resulting in mild flu-like symptoms in the majority of patients, while only a small group of patients develop (bilateral) pneumonia which can rapidly deteriorate in severe acute respiratory distress syndrome^13^. Despite the lower pathogenicity compared to SARS-CoV and MERS-CoV, the high infection rate of SARS-CoV-2 results in large numbers of hospitalizations and deaths worldwide.

The majority of SARS-CoV-2 vaccines are based on the Spike (S) glycoprotein, a homotrimeric glycoprotein that is present on the surface of all coronaviruses and is the main target of protective antibodies. The S protein plays a pivotal role in viral entry and consists of an S1 subdomain including the receptor binding domain (RBD) and an S2 subdomain containing the fusion peptide^14,15^. The S protein of SARS-CoV-2 is most closely related to the S protein of SARS-CoV (∼75% amino acid sequence identity), shows less similarity to MERS-CoV S protein (∼50% identity) and is more distinct from the S proteins of the seasonal hCoVs (∼25-30% identity)^16,17^. Although this sequence identity is considered low from the perspective of eliciting cross-reactive antibodies by vaccination, several studies observed cross-reactivity of COVID-19 sera against other hCoVs^18–22^. In addition, the isolation and engineering of broader hCoV monoclonal antibodies has been reported^23–25^.

In this study, we identify broad hCoV antibody responses following SARS-CoV-2 infection and vaccination. We identify the S2 subdomain as a prominent target for cross-reactive antibodies and demonstrate that cynomolgus macaques vaccinated with a SARS-CoV-2 S protein nanoparticle vaccine elicit detectable antibody responses to other hCoVs. These results will guide the development of a pan-coronavirus vaccine.

## Results

### Convalescent COVID-19 sera contain robust IgG responses to the SARS-CoV-2 S protein

Sera from 50 PCR confirmed SARS-CoV-2 infected patients, aged 22 to 75 years, who suffered from a range of COVID-19 disease severities were obtained 4-6 weeks after symptom onset (Supplementary Table 1). A custom Luminex assay was used to measure IgG antibody levels to the pre-fusion stabilized trimeric SARS-CoV-2 spike (S) protein in these convalescent COVID-19 sera and compared to pre-pandemic healthy donor sera.

Sera from convalescent COVID-19 patients showed 990-fold higher antibody levels to SARS-CoV-2 S protein compared to pre-pandemic healthy donor sera (median 3.8 log_10_ vs. 0.8 log_10_ Median Fluorescent Intensity (MFI), p<0.0001) (Fig. 1a). This is in line with a positive serological response in all but one of the participants using the commercially available WANTAI SARS-CoV-2 RBD total Immunoglobulin serum ELISA (Fig. 1a and Supplementary Table 1). When subdividing the convalescent patients based on admission status, 4-fold higher antibody levels were observed for hospitalized patients compared to non-hospitalized patients (median 4.1 log_10_ vs. 3.5 log_10_ MFI, p<0.0001). SARS-CoV-2 neutralization capacity was also significantly higher (5-fold) in hospitalized patients (3.6 log_10_ vs. 2.9 log_10_ MFI, p<0.0001) and antibody levels and neutralization capacity were strongly correlated in all patients (Spearman r = 0.735, p<0.0001) (Fig. 1b). An 8-fold higher antibody response to the SARS-CoV-2 S protein was observed in the oldest age group of COVID-19 patients (age > 60) compared to the youngest age group (age < 35) (4.1 log_10_ vs. 3.2 log_10_ MFI, p<0.001), which is in line with age being a risk factor for severe COVID-19 (Fig. 1a)^26^. No difference in antibody levels was observed between sex (p=0.71 Mann-Whitney U test). These findings were confirmed with a principal component analysis, which showed two distinct groups based on hospital admission status, and indicated age as an important contributing variable (Fig. 1c, Supplementary Fig. 1). Overall, these results are consistent with previous studies^27,28,29^.

**Figure 1.**
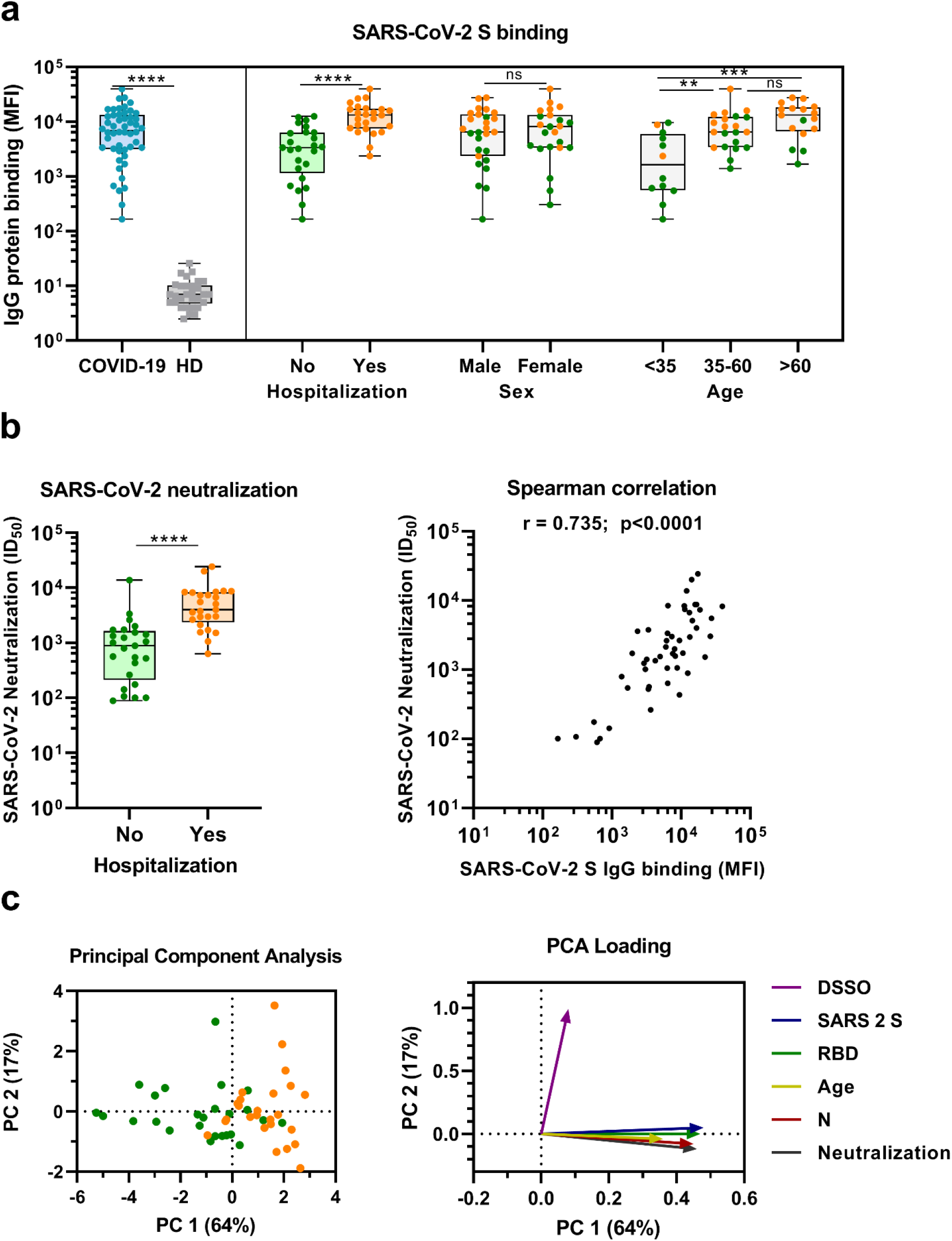
SARS-CoV-2 antibody response in COVID-19 patients. (A) IgG binding of sera to SARS-CoV-2 S protein measured with a custom Luminex assay. Convalescent COVID-19 sera (COVID-19, blue dots, n = 50) were compared to sera of pre-pandemic healthy donors (HD, grey squares, n = 30) using a Mann-Whitney U test. The cohort of COVID-19 patients was subdivided according to admission status, sex and age. Admission status is indicated with orange dots for hospitalized patients (Yes) and green dots if no admission to hospital was needed (No). (B) SARS-CoV-2 pseudovirus neutralization capacity compared between hospitalized patients (orange dots) and patients that did not need admission to hospital (green dots) (left). The correlation between the SARS-CoV-2 pseudovirus neutralization and SARS-CoV-2 S antibody levels in the Luminex assay was determined using Spearman correlation (right). (C) A Principal Component Analysis (PCA) was performed using the following variables: IgG antibodies binding to SARS-CoV-2 S protein (S), RBD and nucleocapsid protein (N), SARS-CoV-2 neutralization, age and days since symptom onset (DSSO). Admission status is indicated with orange dots for hospitalized patients and green dots if no admission to hospital was needed (left). Loading of the principal component plot by each variable is indicated with colored arrows to visualize the contribution of each variable on Principal component (PC) 1 and 2 (right). ns: not significant; ** = p<0.01; *** = p<0.001; **** = p<0.0001. MFI = Median Fluorescent Intensity. S = spike protein. ID_50_ = serum dilution at which 50% of pseudovirus is neutralized. r = Spearman’s rank correlation coefficient.

### Convalescent COVID-19 sera are cross-reactive with all hCoV S proteins

Next, we measured IgG binding to the S proteins of other hCoVs: SARS-CoV, MERS-CoV, hCoV-OC43, hCoV-HKU1, hCoV-229E and hCoV-NL63. We found significantly higher levels of antibodies binding to all hCoV S proteins in convalescent COVID-19 sera compared to pre-pandemic healthy donor sera, with the largest difference for SARS-CoV and MERS-CoV S protein (123 and 11-fold difference between medians, respectively) (Fig. 2a). For the circulating seasonal hCoVs, a 2 to 4-fold difference in median levels of antibodies binding to the S proteins was observed; hCoV-OC43 (4.2-fold, p<0.0001), hCoV-HKU1 (4.3-fold, p<0.0001), hCoV-229E (3.8-fold, p<0.0001) and hCoV-NL63 (1.8-fold, p<0.01). No such difference was found for the control protein Tetanus toxoid (0.8-fold, p=0.85) (Supplementary Fig. 2a).

**Figure 2.**
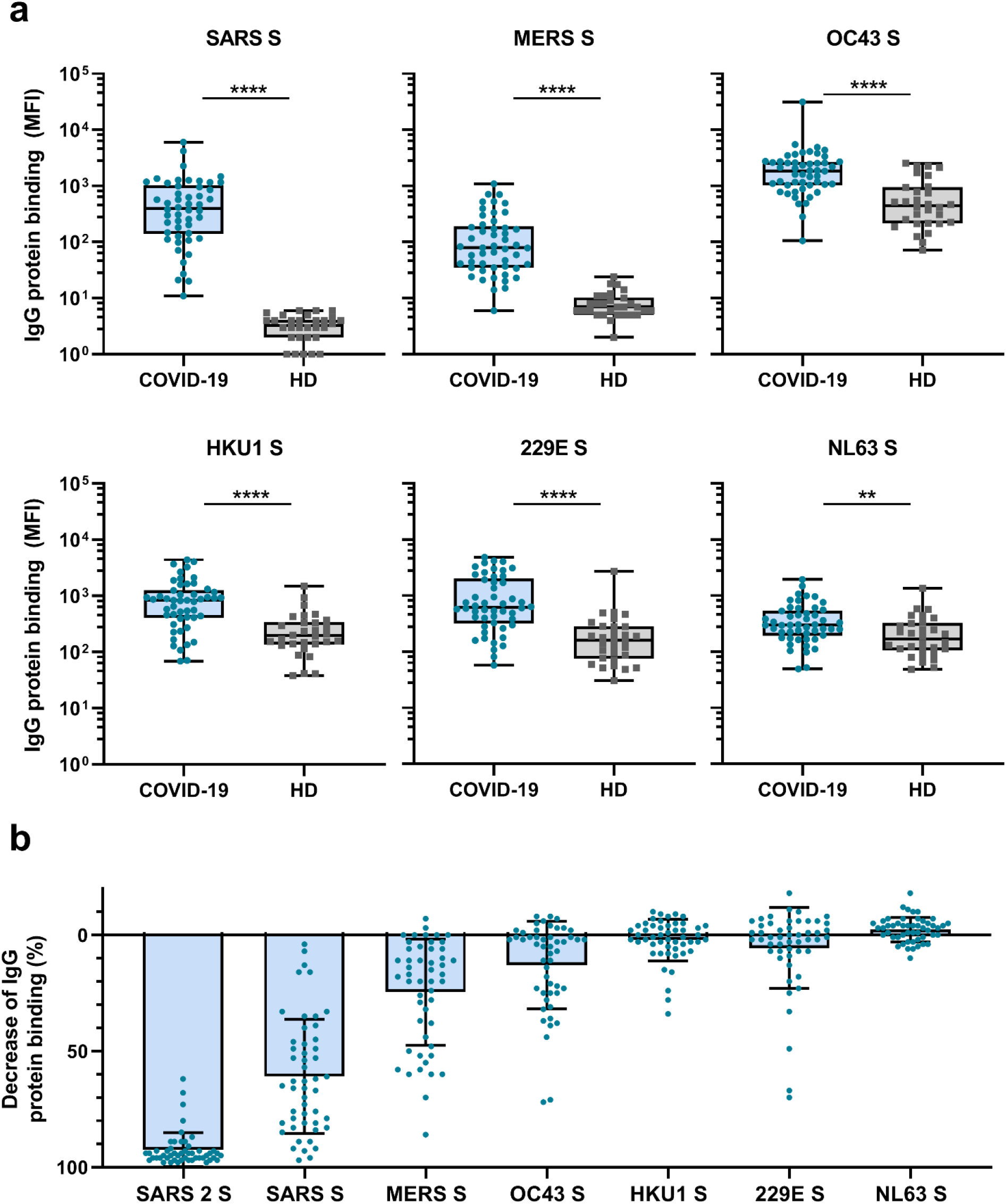
Cross-reactivity to hCoV S proteins in convalescent COVID-19 sera. (A) IgG binding to all hCoV S proteins measured with a custom Luminex assay in convalescent COVID-19 sera (COVID-19, blue dots, n = 50) were compared to pre-pandemic sera from healthy donors (HD, grey squares, n = 30) using a Mann-Whitney U test. Box plots range the minimum and maximum values. ** = p<0.01; **** = p<0.0001. Serum IgG binding to the Tetanus toxoid control protein is shown in Supplementary Fig. 2a. (B) Percent decrease of IgG binding to all other hCoV S proteins in COVID-19 patient sera (n = 50) after depletion with soluble recombinant SARS-CoV-2 S protein. Bars represent the mean decrease of binding IgG as percentage of the total binding IgG observed in undepleted sera and error bars represent the standard deviation. Dots represent the percent decrease of binding IgG observed in individual sera. The percent decrease of IgG binding to Tetanus toxoid control protein in patient sera and healthy donor sera after SARS-CoV-2 S protein depletion is shown in Supplementary Fig. 2b and c and depletion of hCoV S IgG binding in healthy donor sera is shown in Supplementary Fig. 2d. All MFI values are shown in Supplementary Fig. 3a and 6a. MFI = Median Fluorescent Intensity, S = spike protein.

To further explore the specificity of the cross-reactive antibodies in convalescent COVID-19 sera, we performed a depletion with soluble SARS-CoV-2 S protein (Fig. 2b and Supplementary Fig. 3). Depletion of convalescent COVID-19 sera resulted in a 92% reduction of antibody levels against the SARS-CoV-2 S protein. The reduction of antibodies binding to the other hCoV S proteins was strongest for SARS-CoV (61% mean reduction) and MERS-CoV (25%). A more modest reduction of signal was observed for hCoV-OC43 (13% mean reduction) and hCoV-229E S protein (6%) and we observed no depletion of antibodies binding to hCoV-HKU1 (2%) and hCoV-NL63 (−2%). On the individual level, some patients showed up to 72%, 70% and 34% reduction of antibodies binding to hCoV-OC43, hCoV-229E and hCoV-HKU1 S protein after depletion, respectively. No reduction in signal was observed for the control protein Tetanus toxoid after SARS-CoV-2 S depletion (Fig. 2b, Supplementary Fig. 2b). Antibodies binding to other hCoV S proteins in pre-pandemic healthy donor sera did not decrease after depletion (Supplementary Fig. 2c). These depletion experiments corroborate the finding that convalescent COVID-19 sera contain cross-reactive antibodies able to recognize other hCoV S proteins. We observed the highest cross-reactivity for the more closely related hCoVs (Supplementary Table 3). Indeed, the level of cross-reactivity expressed as the reduction of binding antibodies after depletion by SARS-CoV-2 S protein was associated with the level of sequence identity (Spearman r=0.83, p=0.06) (Supplementary Fig. 4).

To further explore the possible functionality of these cross-reactive antibodies, we investigated the correlation between hCoV S protein binding antibodies and SARS-CoV-2 neutralization titers. In addition to the observed correlation between SARS-CoV-2 S protein antibody levels and SARS-CoV-2 neutralization, we found a moderate correlation between SARS-CoV-2 neutralization and antibodies binding to SARS-CoV S protein (r = 0.545, P<0.0001), a weak correlation with antibodies that bind to MERS-CoV, hCoV-HKU1 and hCoV-229E S protein (r = 0.324, 0.308, 0.320 respectively, all p <0.05) and no correlation with antibodies binding to hCoV-OC43 and hCoV-NL63 S protein (r = 0.188 and -0.095, p >0.05) (Supplementary Fig. 6). These correlations suggest that cross-reactive antibodies form a minor component of the neutralizing antibodies induced by SARS-CoV-2 infection. We also investigated if there was an association between cross-reactivity and disease severity with a principal component analysis. Antibodies binding to other hCoV S proteins did not contribute to the separation between patients based on admission status as clearly as was seen for SARS-CoV-2 S protein antibody levels, suggesting a limited impact of cross-reactive antibodies on disease severity (Supplementary Fig. 5).

### Cross-reactive antibodies preferentially target the S2 subdomain

To determine the main target on the SARS-CoV-2 S protein for cross-reactive antibodies, we performed a depletion experiment in which we compared the reduction of IgG binding to hCoV S proteins after depleting the sera with soluble SARS-CoV-2 S1 or S2 subdomains. After depletion with the SARS-CoV-2 S2 subdomain, a 48%, 20% and 19% mean reduction of signal was observed for antibodies binding to SARS-CoV, MERS-CoV, and hCoV-OC43 S protein, respectively, while no reduction was observed after S1 depletion (Fig. 3). Absolute MFI values before and after depletion are shown in Supplementary Fig. 7a. For some individuals, depletion rates reached up to 83% for SARS-CoV, 75% for MERS-CoV and 80% for hCoV-OC43 S protein while there were no individuals with substantial reduction of cross-reactive antibodies after S1 depletion. We did not observe significant depletion of antibodies binding to hCoV-HKU1 and hCoV-229E S proteins by S1 or S2 subdomains of SARS-CoV-2 at the group level. However, in some individuals the signal was reduced by up to 59% for hCoV-HKU1 and 71% for hCoV-229E S protein following depletion with the SARS-CoV-2 S2 subdomain, while such effects were not observed after S1 subdomain depletion. Changes in antibodies that bind to hCoV-NL63 S protein after depletion with SARS-CoV-2 S1 and S2 subdomains were comparable to those observed with the control protein Tetanus toxoid and to the reduction in signal for hCoV S in healthy donors (Supplementary Fig. 3 and 7). SARS-CoV-2 S2 subdomain depletion was significantly correlated with S2 sequence identity (r = 0.90, p = 0.03) while there was no correlation for SARS-CoV-2 S1 subdomain depletion (r = 0.23, p = 0.67) (Supplementary Fig. 4). These results indicate that S2-specific antibodies have a greater contribution to cross-reactivity than S1-specific antibodies, which is in accordance with the higher conservation of the S2 subdomain compared to the S1 subdomain (Supplementary Table 2)^30^.

**Figure 3.**
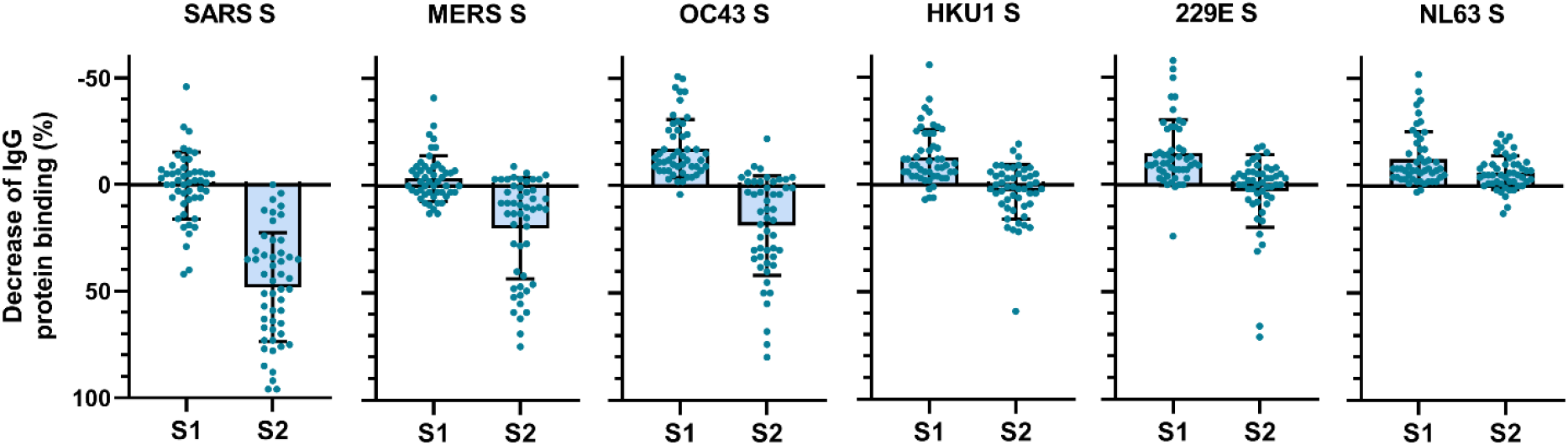
Depletion of S1 and S2 subdomain-specific cross-reactivity in convalescent COVID-19 sera. Percent decrease of IgG binding to all other hCoV S proteins in convalescent COVID-19 sera (n = 50) after depletion with soluble recombinant SARS-CoV-2 S1 or S2 subdomains. Bars represent the mean decrease of binding IgG as percentage of the total binding IgG observed in undepleted sera and error bars represent the standard deviation. Dots represent the percent decrease of binding IgG observed in individual sera. Percent decrease of IgG binding to Tetanus toxoid control protein in patient sera after SARS-CoV-2 S1 and S2 depletion is shown in Supplementary Fig 3a. The percent decrease of IgG binding to hCoV S and Tetanus toxoid protein in healthy donor sera after SARS-CoV-2 S1 and S2 depletion is shown in Supplementary Fig. 7b and c. All MFI values are shown in Supplementary Fig. 3a and 7a. MFI = Median Fluorescent Intensity, S = spike protein.

### SARS-CoV-2 S protein vaccination elicits hCoV cross-reactive antibodies in macaques

The observed cross-reactive antibody response following natural infection with SARS-CoV-2 does not reveal whether these might be *de novo* responses or the result of cross-boosted pre-existing memory from previous hCoV infections. To investigate the ability of SARS-CoV-2 vaccination to induce *de novo* cross-reactive antibodies against hCoVs, we investigated the serum of six cynomolgus macaques which were vaccinated three times with pre-fusion stabilized trimeric SARS-CoV-2 S protein coupled to I53-50 nanoparticles^31^. The vaccination response was studied by comparing serum IgG binding to hCoV S proteins after three vaccinations (week 12) to the pre-immunization baseline (week 0). We observed that all macaques developed detectable antibodies that bind to all hCoV S proteins. Antibodies binding to SARS-CoV, MERS-CoV and hCoV-OC43 S proteins were increased compared to baseline by 3313-, 168-, and 106-fold, respectively (Fig. 4a). This cross-reactivity was already detectable two weeks after the first immunization (Supplementary Fig. 8a). The induction of antibodies that bind to hCoV-HKU1 and hCoV-229E S proteins was relatively low with a 39- and 16-fold increase from baseline, respectively. Antibodies binding to hCoV-NL63 S protein showed only a 6-fold increase from baseline and there was no increase observed for Tetanus toxoid (Fig. 4a and Supplementary Fig. 8b and c). All comparisons were statistically significant except for the Tetanus toxoid. Using the SARS-CoV-2 S protein depletion assay, a 79%, 98% and 99% mean reduction of antibodies binding to SARS-CoV, MERS-CoV and hCoV-OC43 S protein, respectively, was observed (Fig. 4b). In addition, depletion also resulted in a mean reduction of antibodies binding to proteins hCoV-229E S (92%), hCoV-HKU1 S (92%) and hCoV-NL63 S (75%). Virtually no depletion (3%) was observed for the control protein Tetanus toxoid (Supplementary Fig. 9). Together, these results demonstrate that the SARS-CoV-2 S protein is capable of inducing cross-reactive antibodies to both alpha and beta hCoVs, in macaques that we assume have no pre-existing hCoV immunity.

**Figure 4.**
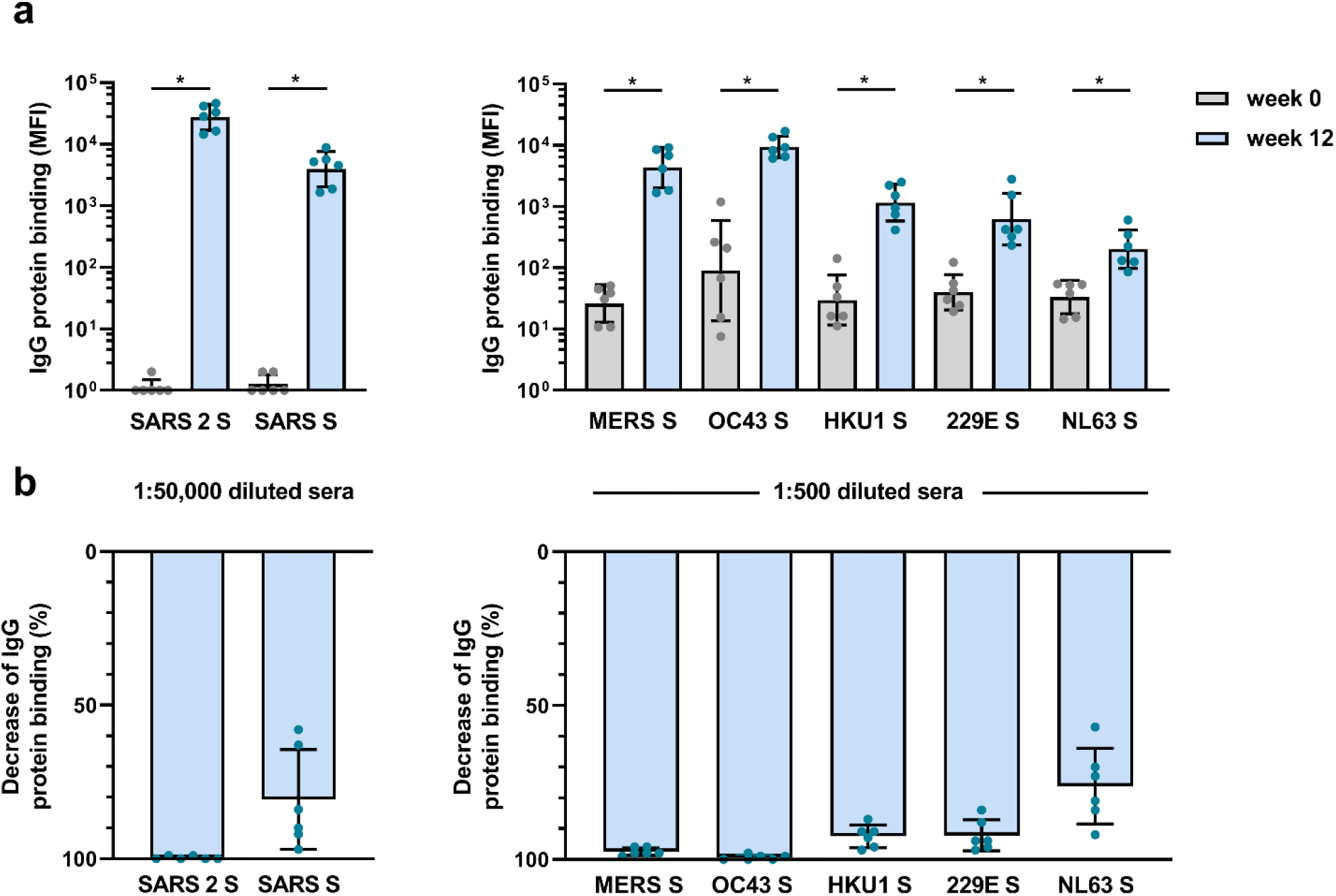
Cross-reactivity and depletion of cross-reactivity to hCoV S proteins in immunized macaques. (A) SARS-CoV-2 S protein specific IgG binding and cross-reactive IgG binding to SARS-CoV S protein at week 0 (pre-immunization baseline) and week 12 (after a total of three immunizations), measured with a custom Luminex assay in 1:50,000 diluted serum of six cynomolgus macaques immunized with a SARS-CoV-2 S nanoparticle vaccine (left) and cross-reactive IgG binding to all other hCoV S proteins, measured in 1:500 diluted serum in the same animals (right). Bars represent the geometric mean of the six animals and error bars the geometric standard deviation; dots represent the individual animals. IgG binding to hCoV S and Tetanus toxoid control protein at weeks 2-12 is shown in Supplementary Fig. 7a and b and IgG binding to Tetanus toxoid control protein at weeks 0 and 12 to is shown in Supplementary Fig. 7c. * = p<0.05 (B) Percent decrease of IgG binding to all hCoV S proteins after depletion with soluble recombinant SARS-CoV-2 S protein. Bars represent the mean decrease of binding IgG as percentage of the total binding IgG observed in undepleted sera of the six cynomolgus macaques at week 12 and error bars represent the standard deviation. Dots represent the percent decrease of binding IgG observed in individual sera. The percent decrease of IgG binding to Tetanus toxoid control protein is shown in Supplementary Fig. 9b and all MFI values are shown in Supplementary Fig. 9a. MFI = Median Fluorescent Intensity, S = spike protein.

## Discussion

As mass vaccination campaigns take place around the globe, there is optimism that the SARS-CoV-2 pandemic will be subdued. However, there will likely be future challenges with emerging SARS-CoV-2 variants as well as with entirely new hCoVs. A number of studies have shown that neutralizing antibodies induced by COVID-19 vaccines have substantially reduced activity against some SARS-CoV-2 variants^3,32,33,34^. Therefore, development of broader SARS-CoV-2 vaccines is needed, with a pan-coronavirus vaccine as the ultimate goal.

In this study, we show that convalescent COVID-19 sera have higher levels of antibodies that bind to other hCoV S proteins compared to pre-pandemic healthy donors. We confirmed this cross-reactivity by showing that SARS-CoV-2 S protein could partially deplete antibodies binding to all hCoV S proteins except hCoV-NL63 in COVID-19 patient sera, while no effect of SARS-CoV-2 S protein depletion was observed in healthy donor sera. Additionally, we see that the elevation of hCoV S antibody levels and the capacity of SARS-CoV-2 S protein to deplete these antibodies is highest for hCoV S proteins that share most sequence homology with the SARS-CoV-2 S protein. Cross-reactive antibodies can be the result of *de novo* responses or due to cross-boosting of pre-existing hCoV immunity. Others have shown cross-reactive antibodies resulting from boosting of pre-existing immunity after natural infection in COVID-19 patients^22,35^. We further explored the possibility to induce *de novo* cross-reactive antibodies by vaccination in a cynomolgus macaque model. These macaques are assumed naïve for hCoVs although infections of macaques have been described^36^. Because of low baseline antibody levels, we expect no previous exposure and therefore solely a *de novo* antibody response following vaccination. All SARS-CoV-2 S protein nanoparticle immunized macaques showed detectable antibodies binding to all hCoV S proteins which could also be depleted with SARS-CoV-2 S protein. These responses after immunization with a vaccine containing a similar pre-fusion stabilized SARS-CoV-2 S protein as many current COVID-19 vaccines, demonstrate that these narrow focused vaccines might already induce low levels of cross-reactive antibodies. However, the strength of the response should be an important consideration, as low levels of antibodies to viral antigens and/or the presence of predominantly non-neutralizing antibodies could potentially lead to Antibody Dependent Enhancement (ADE) of infection^37^. Currently, ADE has not been reported widely for SARS-CoV-2^38^ and for other hCoVs there is no consensus^39^. In addition, it remains unclear in clinical studies if pre-existing immunity against other hCoVs influence the severity of COVID-19^18,20,35,40^. In these 50 COVID-19 patients, we did not find evidence of a clear association between the presence of cross-reactive antibodies and COVID-19 disease severity.

As we show that most of the cross-reactivity is targeting the S2 subdomain of the S protein, we propose that this subdomain should receive more attention in vaccine design. Interestingly, although most neutralizing antibodies target the S1 subdomain including the RBD, protective neutralizing antibodies targeting the S2 subdomain have also been identified^41^. Furthermore, analogous to some influenza virus hemagglutinin stem antibodies^42^, antibody effector functions might also contribute to protective immunity by S2 antibodies^43^. Additional possibilities to improve breadth include the use of consensus or mosaic vaccine designs similar to what has been done in HIV-1 vaccine research^44,45^, or the inclusion of an S2-based booster vaccine after SARS-CoV-2 vaccination to amplify and broaden the response.

The results from this study on the presence and specificity of cross-reactive antibodies to other hCoVs after SARS-CoV-2 infection and vaccination, emphasize the feasibility of broad coronavirus vaccines and may guide future vaccine designs.

## Materials & methods

### Study population

Sera of 50 SARS-CoV-2 infected adults were collected 4-6 weeks after symptom onset through the cross-sectional COVID-19 Specific Antibodies (COSCA) study (NL 73281.018.20) as described previously^46^. In short, all participants had at least one nasopharyngeal or oropharyngeal swab positive for SARS-CoV-2 as determined by qRT-PCR (Roche LightCycler480, targeting the Envelope-gene 113bp) and 49 of 50 participants were serological positive for SARS-CoV-2 (Wantai, WS-1096, total SARS-CoV-2 RBD targeting antibodies). Patient demographics and medical history were collected and used to score the maximum disease severity by using the WHO disease severity criteria of COVID-19 (https://www.who.int/publications/i/item/clinical-management-of-covid-19, accessed on 16-10-2020). The COSCA study was conducted at the Amsterdam University Medical Centre, location AMC, the Netherlands and approved by the local ethical committee (NL 73281.018.20). All individuals provided written informed consent before participating.

Sera of 30 healthy donors were kindly provided by the Dutch Institute for Public Health and Environment (RIVM). Sera donated in 2019 were used to exclude possible exposure to SARS-CoV-2. No seasonal influence was observed when analyzing IgG binding to the seasonal coronaviruses of control sera obtained at different calendar months (data not shown).

### Cynomolgus Macaques

Cynomolgus macaques (n = 6) received 50 µg of SARS-CoV-2 S-I53-50NP adjuvanted with 500 µg of MPLA liposomes (Polymun Scientific, Klosterneuburg, Austria) diluted in PBS by intramuscular route at weeks 0, 4 and 10. Blood samples were taken at baseline and weeks 2, 4, 6, 8, 10 and 12 after vaccination. Additional information is described by Brouwer et al. (2021)^31^.

### Protein designs

A prefusion stabilized S protein ectodomain of SARS-CoV-2 and SARS-CoV with a T4 trimerization domain and hexahistidine (His) tag were previously described^46^. A prefusion stabilized S protein ectodomains construct of hCoV-HKU1 was designed as described previously^47^. Specifically, a gene encoding amino acids 1-1276, in which the furin cleavage site (amino acids 752-756) was replaced with a GGSGS sequence and amino acids at position 1067 and 1068 were mutated to prolines, was ordered and cloned into a pPPI4 plasmid containing a T4 trimerization domain followed by a hexahistidine tag. The same procedure was used to generate prefusion stabilized S proteins of MERS-CoV, hCoV-229E, hCoV-NL63, and hCoV-OC43. For the prefusion stabilized S protein ectodomain of hCoV-229E, a gene encoding amino acids 1-1082, with proline substitutions at positions 871 and 872, was ordered. The prefusion stabilized S protein ectodomain construct of hCoV-NL63 consisted of amino acids 1-1263 and introduction of proline substitutions at positions 1052 and 1053. For MERS-CoV, a gene encoding amino acids 1-1214 with proline substitutions at positions 1060 and 1061, and the furin cleavage site (amino acids 748-751) replaced with an ASVG sequence. The prefusion stabilized S protein ectodomain construct of hCoV-OC43 was designed as described previously^48^. Specifically, this construct consisted of amino acids 1-1263, which had the furin cleavage site replaced with a GGSGG sequence and proline substitutions at positions 1070 and 1071. GenBank ID MN908947.3 (SARS-CoV-2) ABD72984.1 (SARS-CoV), AHI48550.1 (MERS-CoV), AAT84362.1 (hCoV-OC43), Q0ZME7 (hCoV-HKU1), NP_073551.1 (hCoV-229E) and AKT07952.1 (hCoV-NL63) served as templates for the protein designs.

### Protein expression and purification

The Tetanus toxoid protein was acquired from Creative Biolabs. SARS-CoV-2 Nucleocapsid protein was kindly provided by Gestur Vidarsson and Federica Linty of Sanquin Research, Amsterdam, the Netherlands. All other proteins were produced in HEK293F cells (Invitrogen) maintained in Freestyle medium (Life Technologies). Transfections were performed using Polyethylenimine Hydrochloride (PEI) MAX (Polysciences) at 1 mg/L and the expression plasmids at 312.5 µg/L in a 3:1 ratio in 50 mL OptiMEM (Gibco) per L. Supernatants were harvested seven days post transfection by centrifugation at 4,000 rpm for 30 min followed by filtration of the supernatant using 0.22 µM Steritop filter units (Merck Millipore). The His-tagged proteins were purified from the clarified supernatant with affinity chromatography using NiNTA agarose beads (Qiagen). Eluates were concentrated and buffer exchanged to PBS using 100 kDa molecular weight cut-off (MWCO) Vivaspin centrifugal concentrators. Further purification to remove aggregated and monomeric protein fractions was performed using Size Exclusion Chromatography on a Superose6 increase 10/300 GL column (GE Healthcare) using PBS as buffer. Trimeric S proteins eluted at a volume of approximately 13 mL. Fractions containing trimeric protein were pooled and concentrated using 100 kDa MWCO Vivaspin centrifugal concentrators. Resulting protein concentrations were determined using a Nanodrop 2000 Spectrophotometer and proteins were stored at -80°C until needed.

### Protein coupling to Luminex beads

Proteins were covalently coupled to Luminex Magplex beads using a two-step carbodiimide reaction and a ratio of 75 µg protein to 12.5 million beads for SARS-CoV-2 S protein. Other proteins were coupled equimolar to SARS-CoV-2 S protein. Luminex Magplex beads (Luminex) were washed with 100 mM monobasic sodium phosphate pH 6.2 and activated by addition of Sulfo-N-Hydroxysulfosuccinimide (Thermo Fisher Scientific) and 1-Ethyl-3-(3-dimethylaminopropyl) carbodiimide (Thermo Fisher Scientific) and incubated for 30 min on a rotator at room temperature. The activated beads were washed three times with 50 mM MES pH 5.0 and the proteins were diluted in 50 mM MES pH 5.0 and added to the beads. The beads and proteins were incubated for three h on a rotator at room temperature. Afterwards, the beads were washed with PBS and blocked with PBS containing 2% BSA, 3% Fetal calf serum and 0.02% Tween-20 at pH 7.0 for 30 min on a rotator at room temperature. Finally, the beads were washed and stored at 4°C in PBS containing 0.05% Sodium Azide and used within six months. Detection of the His tag on each S protein-coupled bead was used to confirm the amount of protein on the beads.

### Luminex assays

Optimization experiments determined the optimal concentration for detection of hCoV S protein binding antibodies to be 1:10,000 dilution for COVID-19 patients, 1:50,000 for SARS-CoV and SARS-CoV-2 detection in cynomolgus macaques and 1:500 for detection of binding antibodies to other hCoV S proteins in cynomolgus macaques. 50 µL of a working bead mixture containing 20 beads per µl of each region was incubated overnight with 50 µl of diluted serum. Plates were sealed and incubated on a plate shaker overnight at 4°C. The next day, plates were washed with TBS containing 0.05% Tween-20 (TBST) using a hand-held magnetic separator. Beads were resuspended in 50 µl of Goat-anti-human IgG-PE (Southern Biotech) and incubated on a plate shaker at room temperature for 2 h. Afterwards, the beads were washed with TBST and resuspended in 70 µl Magpix drive fluid (Luminex). The beads were agitated for a few min on a plate shaker at room temperature and then read-out was performed on a Magpix (Luminex). Resulting MFI values are the median of approximately 50 beads per well and were corrected by subtraction of MFI values from buffer and beads only wells. A titration of serum of one convalescent COVID-19 patient as well as positive and negative controls were included on each plate to confirm assay performance. Assays were performed twice with similar results (Supplementary Fig. 9).

### Depletion Luminex assay

Convalescent COVID-19 patient sera were diluted 1:10,000 with addition of 10 µg/mL soluble recombinant SARS-CoV-2 pre-fusion stabilized S protein, S1 or S2 subdomain (ABclonal Biotechnology) or without protein (as undepleted controls). Immunized macaque sera were diluted 1:500 with addition of 30 µg/mL SARS-CoV-2 S protein. Incubation was performed in uncoated 96-wells plates on a shaker at room temperature for 1 h. Then, 50 µl of a working bead mixture containing 20 beads per µl of each region was added and the plates were incubated 2 h on a shaker at room temperature. After 2 h, the above described Luminex protocol was continued starting with the TBST washes and the Goat-anti-human IgG-PE incubation.

### SARS-CoV-2 pseudovirus neutralization assay

The pseudovirus neutralization assay was performed as described previously^31^. Briefly, HEK293T cells expressing the SARS-CoV-2 receptor ACE2 were seeded in poly-L-lysine coated 96-wells plates and the next day triplicate serial dilutions of heat-inactivated serum samples were prepared, mixed 1:1 with SARS-CoV-2 pseudovirus, incubated for 1 h at 37°C and then added in a 1:1 ratio to the cells. After 48 h, the cells were lysed, transferred to half-area 96-wells white microplates (Greiner Bio-One) and Luciferase activity was measured using the Nano-Glo Luciferase Assay System (Promega) with a Glomax system (Turner Biosystems). Relative luminescence units were normalized to the units from cells infected with pseudovirus in absence of serum. Neutralization titers (ID_50_) were the serum dilution at which infectivity was inhibited 50%.

### Statistical Analysis

Luminex data were log transformed prior to any statistical analysis. P values below 0.05 were considered statistically significant. Mann-Whitney U-tests for unpaired comparisons, Wilcoxon matched-pairs signed rank test for paired comparisons and Spearman correlations were performed in GraphPad Prism 8.3.0. Principal component analysis was performed in Matlab 9.6 (R2019a).

## Data Availability

Data supporting the findings in this manuscript are available from the corresponding author upon request.

## Acknowledgements

We are thankful to the participants of the COSCA-study for their contribution to this research. In addition, we thank the research team of the RECoVERD/VIS-cohort study for assisting with the recruitment and inclusion of participants. We thank Gestur Vidarsson and Federica Linty of Sanquin, Amsterdam, The Netherlands for providing the SARS-CoV-2 Nucleocapsid protein. We thank Mathieu A. F. Claireaux, Aafke Aartse, Ronald Derking and Jonne L. Snitselaar for providing assay controls and Aafke Aartse for manuscript editing.

This work was supported by a Netherlands Organization for Scientific Research (NWO) Vici grant (to R.W.S.); by the Bill & Melinda Gates Foundation through the Collaboration for AIDS Vaccine Discovery (CAVD) grants OPP1111923, OPP1132237, and INV-002022 to R.W.S.; by Health Holland PPS-allowance LSHM20040 (to M.J.v.G.) and by ZonMw (to M.D.d.J.). M.J.v.G. is a recipient of an AMC Fellowship from Amsterdam UMC and a COVID-19 grant from the Amsterdam Institute for Infection and Immunity. R.W.S and M.J.v.G. are recipients of support from the University of Amsterdam Proof of Concept fund (contract no. 200421) as managed by Innovation Exchange Amsterdam (IXA). A.W.C is supported by an NHMRC career development fellowship. The funders had no role in study design, data collection, data analysis, data interpretation, or data reporting.

## Author contributions

Conceived and designed the experiments: M.G., K.v.d.S., A.W.C., R.W.S., M.J.v.G. Arranged medical ethical approval, recruitment of study participants, and collection of study material: K.v.d.S, H.D.G.v.W., E.W., B.J.V., O.J.A.F., P.J.d.V., T.M.B., M.D.d.J, M.Pr., G.J.d.B.

Performed the experiments: M.G., K.v.d.S., J.A.B, M.Po., M.O.

Provided reagents: P.J.M.B., M.B., P.M., N.D.B., R.L.G., D.E., T.P.L.B.

Analyzed and interpreted the data: M.G., K.v.d.S., A.W.C., M.J.v.G.

Wrote the manuscript: M.G., K.v.d.S., R.W.S., M.J.v.G.

Edited the manuscript: all

## Competing interests

The authors declare no competing interests.

## Supplementary figure legends

**Supplementary Figure 1. Principal Component Analysis showing the influence of sex and disease severity on the SARS-CoV-2 IgG response**.

A Principal Component Analysis was performed using the following variables: IgG antibodies to SARS-CoV-2 S protein (S), RBD and nucleocapsid protein (N), SARS-CoV-2 neutralization, age and days since symptom onset (DSSO). Patients were subdivided based on sex (A) and WHO severity score (B). A description of the WHO severity scoring is found in Supplementary Table 1. The loading of the principal component plot by each variable is shown in Fig. 1.

**Supplementary Figure 2. Antibody reactivity to Tetanus toxoid in convalescent COVID-19 sera and depletion of antibodies binding to all proteins in healthy donors**.

(A) IgG binding to Tetanus toxoid protein measured with a custom Luminex assay in sera of convalescent COVID-19 sera (COVID-19, orange dots, n = 50) compared to sera of pre-pandemic healthy donors (HD, grey squares, n = 30) using a Mann-Whitney U test. Box plots range the minimum and maximum values. Ns = non-significant. (B) Percent decrease of IgG binding to Tetanus toxoid protein in COVID-19 patient sera (n = 50) after depletion with soluble recombinant SARS-CoV-2 S protein. (C) Percent decrease of IgG binding to Tetanus toxoid protein after depletion with soluble recombinant SARS-CoV-2 S protein in healthy donors (n = 30) and (D) Percent decrease of IgG binding to hCoV S proteins after depletion with soluble recombinant SARS-CoV-2 S protein in healthy donors (n = 30). Bars represent the mean decrease of binding IgG as percentage of the total binding IgG observed in undepleted sera of all COVID-19 patients (B) or healthy donors (C and D) and error bars represent the standard deviation. Dots represent the percent decrease of binding IgG in individual sera. All MFI values are shown in Supplementary Fig. 3 and 7. MFI = median fluorescent intensity.

**Supplementary Figure 3. Raw data and control data from depletion assays on convalescent COVID-19 sera**.

(A) MFI values of IgG binding to all hCoV S proteins and Tetanus toxoid control protein, measured with a custom Luminex assay in sera of convalescent COVID-19 sera (n = 50) after depletion with soluble recombinant SARS-CoV-2 S protein, S1 subdomain and S2 subdomain. Box plots range the minimum and maximum values. - = undepleted sera, S = trimeric S protein depleted sera, S1 = S1 subdomain depleted sera, S2 = S2 subdomain depleted sera. (B) Percent decrease of IgG binding to Tetanus toxoid control protein in sera of convalescent COVID-19 patients (n = 50) after depletion with soluble recombinant SARS-CoV-2 S1 or S2 subdomain. Bars represent the mean decrease of binding IgG as percentage of the total binding IgG observed in undepleted sera of all COVID-19 patients and error bars represent the standard deviation. Dots represent the percent decrease found in individual patient sera. MFI = median fluorescent intensity, S = spike protein.

**Supplementary Figure 4. Correlation between sequence identity and reduction of cross-reactive antibodies in depletion assay for S, S1 and S2**

The correlation between S protein (a), S1 subdomain (b) and S2 subdomain (c) sequence identity of SARS-CoV-2 with the different hCoV S proteins (also shown in Supplementary Table 2) and the reduction of antibodies binding to hCoV S proteins in the SARS-CoV-2 S protein depletion assay (also shown in Figure 2b) was determined using Spearman correlation. S = spike protein, r = Spearman’s rank correlation coefficient.

**Supplementary Figure 5. Spearman correlations between SARS-CoV-2 neutralization and antibody reactivity to hCoV S and Tetanus toxoid**.

The correlation between the SARS-CoV-2 pseudovirus neutralization (also shown in Fig. 1b) and binding IgG binding measured with a custom Luminex assay (also shown in Fig. 2a) in sera of convalescent COVID-19 patients (n = 50) was determined using Spearman correlation. MFI = median fluorescent intensity, S = spike protein, ID_50_ = serum dilution at which 50% of pseudovirus is neutralized, r = Spearman’s rank correlation coefficient.

**Supplementary Figure 6. Principal Component Analysis including SARS-CoV-2 binding antibodies, neutralization, clinical characteristics and cross-reactivity**. The Principal Component Analysis was performed using the following variables: IgG binding to all hCoV S proteins including SARS-CoV-2, IgG binding to SARS-CoV-2 RBD and nucleocapsid protein (N), SARS-CoV-2 neutralization, patient age and days since symptom onset (DSSO). Color indicates the difference between patients that needed hospitalization (orange) or stayed in home quarantine (green) (top left), between males (grey) and females (purple) (bottom left) and between patients with different WHO severity scores (bottom right, description of scoring is found in Supplementary Table 1). The loading of each variable for Principal Component (PC) 1 and 2 are shown (top right).

**Supplementary Figure 7. Raw data and control data from depletion assays on healthy donor sera**.

(A) MFI values of IgG binding to all hCoV S proteins (top and middle row) and control proteins (bottom row) measured with a custom Luminex assay in sera of pre-pandemic healthy donors (n = 30) after depletion with soluble recombinant SARS-CoV-2 S protein, S1 subdomain and S2 subdomain. Box plots range the minimum and maximum values. - = undepleted sera, S = trimeric S protein depleted sera, S1 = S1 subdomain depleted sera, S2 = S2 subdomain depleted sera. (B) Percent decrease of IgG binding to hCoV S proteins in sera of healthy donors (n = 30) after depletion with soluble recombinant SARS-CoV-2 S1 subdomain or S2 subdomain. (C) Percent decrease of IgG binding to Tetanus toxoid control protein in sera of healthy donors (n = 30) after depletion with soluble recombinant SARS-CoV-2 S1 subdomain or S2 subdomain. Bars represent the mean decrease of binding antibodies as percentage of the total binding IgG observed in undepleted sera of all healthy donors and error bars represent the standard deviation. Dots represent the percent decrease of binding IgG found in individual sera. MFI = median fluorescent intensity, S = spike protein.

**Supplementary Figure 8. Antibodies binding to hCoV S and tetanus toxoid in immunized cynomolgus macaques over time**.

(A) IgG response over time in serum of six cynomolgus macaques immunized at week 0, 4 and 10 with a SARS-CoV-2 Spike nanoparticle vaccine, measured at a 1:50,000 serum dilution for SARS-CoV-2 and SARS-CoV S and at 1:500 serum dilution for MERS-CoV, hCoV-OC43, hCoV-HKU1, hCoV-229E and hCoV-NL63-CoV S. (B) IgG binding to Tetanus toxoid control protein, measured in 1:500 diluted serum. All MFI values are background corrected by subtraction of beads and buffer only wells. (C) IgG binding to Tetanus toxoid control protein at week 0 (pre-immunization baseline) and week 12 (after a total of three immunizations), presented in a bar chart for comparison with Fig. 4a. ns = not significant, MFI = median fluorescent intensity, S = spike protein.

**Supplementary Figure 9. Reactivity and depletion of antibodies to hCoV S proteins and Tetanus Toxoid in immunized cynomolgus macaques**.

(A) MFI values of IgG binding to all hCoV S proteins and Tetanus toxoid control protein with and without depletion with soluble recombinant SARS-CoV-2 S protein, measured by Luminex assay in 1:50,000 diluted serum (for SARS-CoV-2 and SARS-CoV) or 1:500 diluted serum (for all other proteins) for six cynomolgus macaques immunized at week 0, 4 and 10 with a SARS-CoV-2 S protein nanoparticle vaccine. The line indicates the geometric mean. (B) Percent decrease of IgG binding to Tetanus toxoid control protein in 1:500 diluted sera at week 12 after depletion with soluble recombinant SARS-CoV-2 S protein, presented in bar charts for comparison with Fig. 4b. Bars represent the mean decrease of binding IgG as percentage of the total binding IgG observed in undepleted sera of the six cynomolgus macaques. Dots represent the percent decrease of binding IgG in each individual animal. MFI = median fluorescent intensity, S = spike protein.

**Supplementary Figure 10. Reproducibility of the Luminex assay**.

The reproducibility of the custom Luminex assay is shown by plotting data from two independent assays performed on different days with the same samples. The data on the Y axis is found in Fig. 3 and the data on the X axis is also found in Fig. 1 and 2. The quality of the replicates is presented by the R^2^ and slope of a simple linear regression performed on the log_10_ transformed MFI data. MFI = median fluorescent intensity, S = spike protein, N = nucleocapsid, RBD = receptor binding domain.

## Notes

### Competing Interest Statement

The authors have declared no competing interest.

### Clinical Trial

This study is a cross-sectional observational study without intervention, it was not registered with a trial ID.

### Author Declarations

For the convalescent COVID-19 sera, approval was granted by the ethical review board of the Amsterdam University Medical Centers, location AMC, the Netherlands (NL 73281.018.20) and for the healthy controls, anonymized leftover serum from routine diagnostics for which ethical approval was waived by the ethics committee of the National Institute of Public Health and the Environment was used. The study was performed in accordance with the guidelines for sharing of patient data of observational scientific research in emergency situations as issued by the Commission on Codes of Conduct of the Foundation Federation of Dutch Medical Scientific Societies (https://www.federa.org/federa-english).

