## Supplementary Materials for "Cross-reactive antibodies after SARS-CoV-2 infection and vaccination"

**Supplementary Table 1. Sociodemographics, clinical characteristics and severity scoring for COVID-19 patients**

| Sociodemographics and Clinical Characteristics | Number of patients n (%) |
|---|---|
| Sex | |
| Male | 27 (54) |
| Female | 23 (46) |
| Age (years) | |
| <35 | 12 (24) |
| 35-60 | 21 (42) |
| >60 | 17 (34) |
| Hospitalization | |
| Home | 25 (50) |
| Admission | 25 (50) |
| Disease severity | |
| 0 | 0 (0) |
| 1 | 16 (32) |
| 2 | 17 (34) |
| 3 | 14 (28) |
| 4 | 3 (6) |
| Serological positive (WANTAI) | |
| | 49 (98) |

| Score | Short name | Criteria |
|---|---|---|
| 0 | Asymptomatic | SARS-CoV-2 proven infection without developing symptoms of COVID-19 |
| 1 | Mild | Symptoms of COVID-19 without evidence of viral pneumonia or hypoxia |
| 2 | Moderate | Clinical signs of pneumonia but no signs of severe pneumonia, including SpO2 ≥ 90% on room air |
| 3 | Severe | Clinical signs of severe pneumonia: respiratory rate > 30 breaths/min, spO2 <90% on room air |
| 4 | Critical | ICU admission (ARDS, Sepsis, Septic shock) |

**Supplementary Table 2. Sequence identity matrices for the ectodomains of all hCoV S proteins**

## S

| | SARS 2 S | SARS S | MERS S | OC43 S | HKU1 S | 229E S | NL63 S |
|---|---|---|---|---|---|---|---|
| SARS 2 S | 100% | | | | | | |
| SARS S | 75,4% | 100% | | | | | |
| MERS S | 30,9% | 31,4% | 100% | | | | |
| OC43 S | 30,0% | 30,6% | 31,7% | 100% | | | |
| HKU1 S | 30,3% | 31,1% | 32,2% | 65,9% | 100% | | |
| 229E S | 27,4% | 27,2% | 28,4% | 27,7% | 28,8% | 100% | |
| NL63 S | 26,2% | 24,8% | 27,1% | 28,1% | 28,3% | 65,1% | 100% |

## S1

| | SARS 2 S1 | SARS S1 | MERS S1 | OC43 S1 | HKU1 S1 | 229E S1 | NL63 S1 |
|---|---|---|---|---|---|---|---|
| SARS 2 S1 | 100% | | | | | | |
| SARS S1 | 66,8% | 100% | | | | | |
| MERS S1 | 21,0% | 22,1% | 100% | | | | |
| OC43 S1 | 25,1% | 24,7% | 23,4% | 100% | | | |
| HKU1 S1 | 23,9% | 25,2% | 26,4% | 61,5% | 100% | | |
| 229E S1 | 17,5% | 16,9% | 18,1% | 16,5% | 19,4% | 100% | |
| NL63 S1 | 16,4% | 14,6% | 15,1% | 16,9% | 17,5% | 51,7% | 100% |

## S2

| | SARS 2 S2 | SARS S2 | MERS S2 | OC43 S2 | HKU1 S2 | 229E S2 | NL63 S2 |
|---|---|---|---|---|---|---|---|
| SARS 2 S2 | 100% | | | | | | |
| SARS S2 | 87,6% | 100% | | | | | |
| MERS S2 | 44,5% | 43,4% | 100% | | | | |
| OC43 S2 | 41,8% | 40,9% | 45,2% | 100% | | | |
| HKU1 S2 | 41,3% | 40,6% | 43,5% | 71,7% | 100% | | |
| 229E S2 | 38,0% | 37,7% | 38,0% | 35,0% | 35,0% | 100% | |
| NL63 S2 | 38,2% | 35,9% | 36,9% | 36,6% | 35,4% | 77,7% | 100% |

### RBD

| | SARS 2 RBD | SARS RBD | MERS RBD | OC43 RBD | HKU1 RBD | 229E RBD | NL63 RBD |
|---|---|---|---|---|---|---|---|
| SARS 2 RBD | 100% | | | | | | |
| SARS RBD | 73,4% | 100% | | | | | |
| MERS RBD | 19,2% | 21,3% | 100% | | | | |
| OC43 RBD | 23,1% | 24,6% | 27,1% | 100% | | | |
| HKU1 RBD | 19,7% | 23,4% | 27,5% | 60,0% | 100% | | |
| 229E RBD | 16,7% | 12,8% | 23,6% | 17,1% | 21,1% | 100% | |
| NL63 RBD | 12,5% | 17,0% | 20,0% | 17,1% | 15,6% | 57,3% | 100% |

Sequence identity matrices were composed of all coronavirus spike proteins in this study. All sequences comprise only the truncated ectodomain of each spike as was used to generate the recombinant proteins. S1, S2 and RBD were defined as noted in the corresponding Genbank sequences (see methods). Multiple sequence alignments were performed and sequence identities calculated using Clustal Omega 1.2.4.

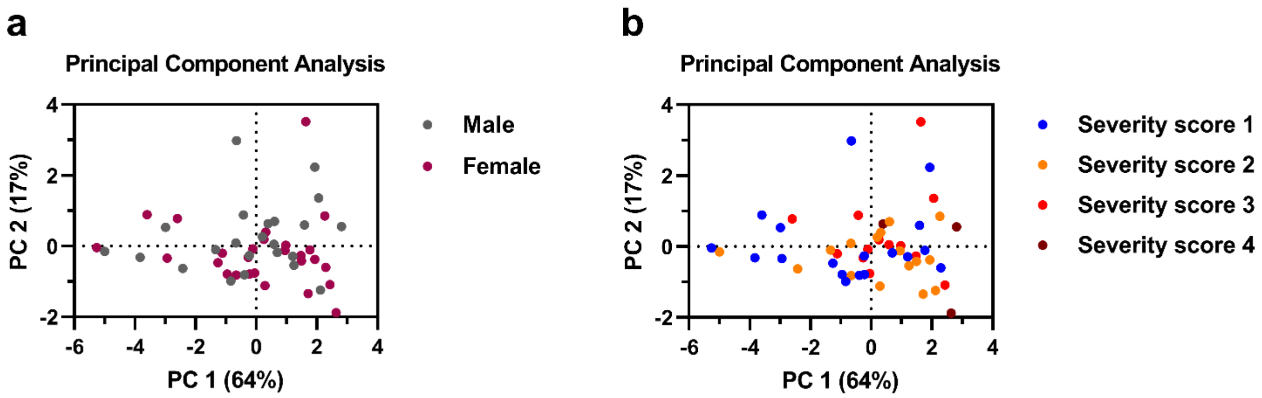


**Supplementary Figure 1. Principal Component Analysis showing the influence of sex and disease severity on the SARS-CoV-2 IgG response.**

A Principal Component Analysis was performed using the following variables: IgG antibodies to SARS-CoV-2 S protein (S), RBD and nucleocapsid protein (N), SARS-CoV-2 neutralization, age and days since symptom onset (DSSO). Patients were subdivided based on sex (A) and WHO severity score (B). A description of the WHO severity scoring is found in Supplementary Table 1. The loading of the principal component plot by each variable is shown in Fig. 1.

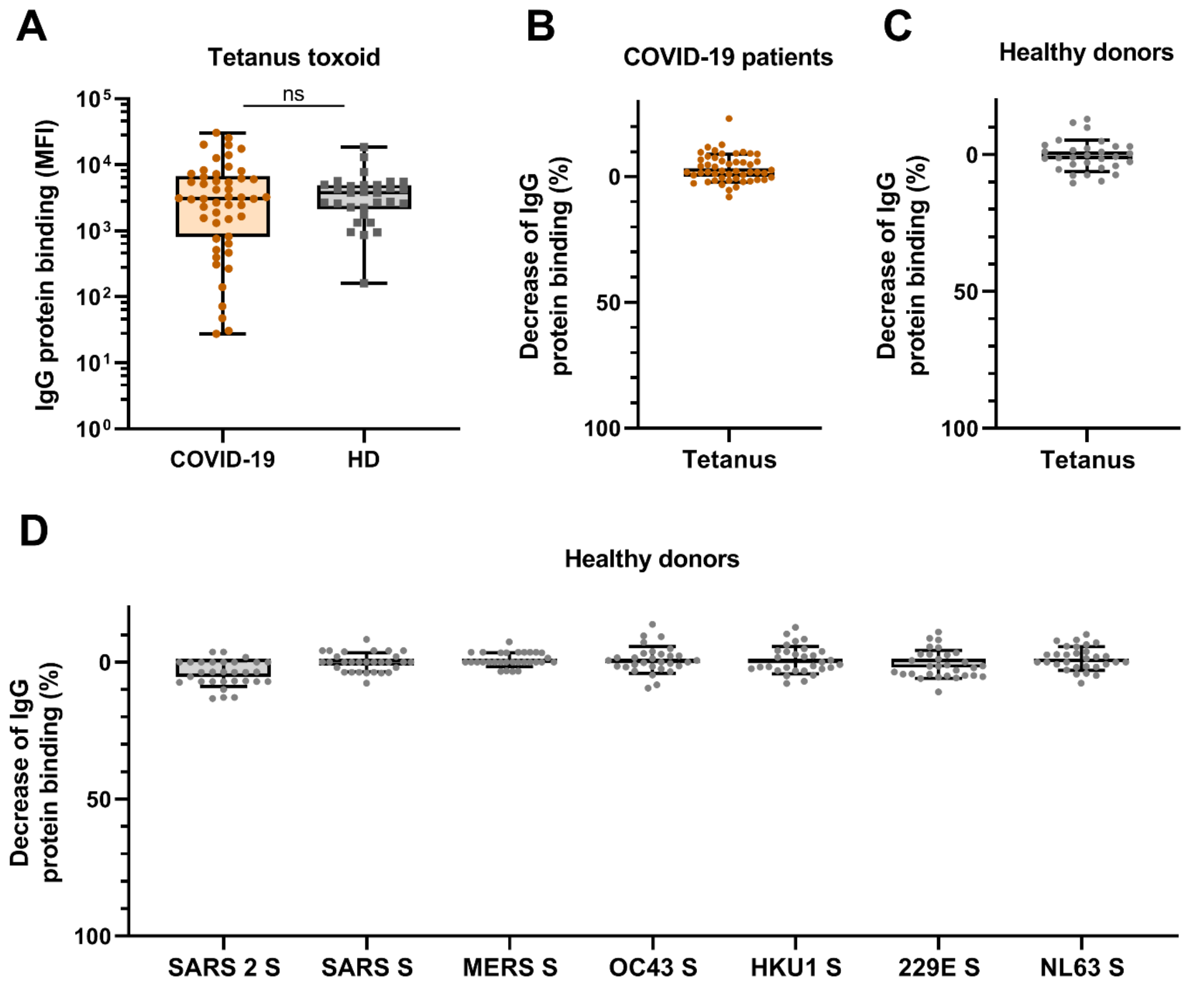


**Supplementary Figure 2. Antibody reactivity to Tetanus toxoid in convalescent COVID-19 sera and depletion of antibodies binding to all proteins in healthy donors.**

(A) IgG binding to Tetanus toxoid protein measured with a custom Luminex assay in sera of convalescent COVID-19 sera (COVID-19, orange dots, n = 50) compared to sera of pre-pandemic healthy donors (HD, grey squares, n = 30) using a Mann-Whitney U test. Box plots range the minimum and maximum values. Ns = non-significant. (B) Percent decrease of IgG binding to Tetanus toxoid protein in COVID-19 patient sera (n = 50) after depletion with soluble recombinant SARS-CoV-2 S protein. (C) Percent decrease of IgG binding to Tetanus toxoid protein after depletion with soluble recombinant SARS-CoV-2 S protein in healthy donors (n = 30) and (D) Percent decrease of IgG binding to hCoV S proteins after depletion with soluble recombinant SARS-CoV-2 S protein in healthy donors (n = 30). Bars represent the mean decrease of binding IgG as percentage of the total binding IgG observed in undepleted sera of all COVID-19 patients (B) or healthy donors (C and D) and error bars represent the standard deviation. Dots represent the percent decrease of binding IgG in individual sera. All MFI values are shown in Supplementary Fig. 3 and 7. MFI = median fluorescent intensity.

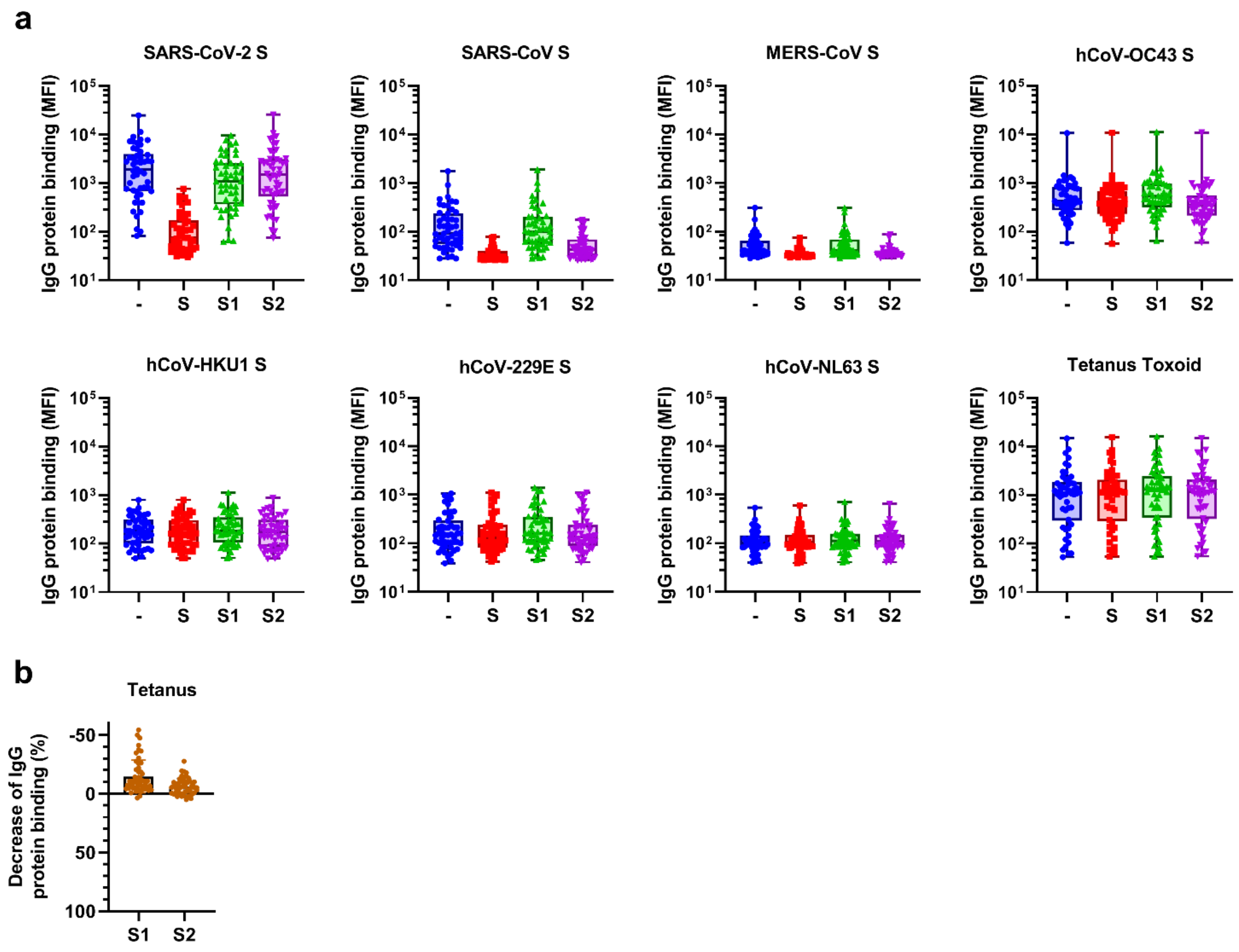


**Supplementary Figure 3. Raw data and control data from depletion assays on convalescent COVID-19 sera.**

(A) MFI values of IgG binding to all hCoV S proteins and Tetanus toxoid control protein, measured with a custom Luminex assay in sera of convalescent COVID-19 sera (n = 50) after depletion with soluble recombinant SARS-CoV-2 S protein, S1 subdomain and S2 subdomain. Box plots range the minimum and maximum values. - = undepleted sera, S = trimeric S protein depleted sera, S1 = S1 subdomain depleted sera, S2 = S2 subdomain depleted sera. (B) Percent decrease of IgG binding to Tetanus toxoid control protein in sera of convalescent COVID-19 patients (n = 50) after depletion with soluble recombinant SARS-CoV-2 S1 or S2 subdomain. Bars represent the mean decrease of binding IgG as percentage of the total binding IgG observed in undepleted sera of all COVID-19 patients and error bars represent the standard deviation. Dots represent the percent decrease found in individual patient sera. MFI = median fluorescent intensity, S = spike protein.

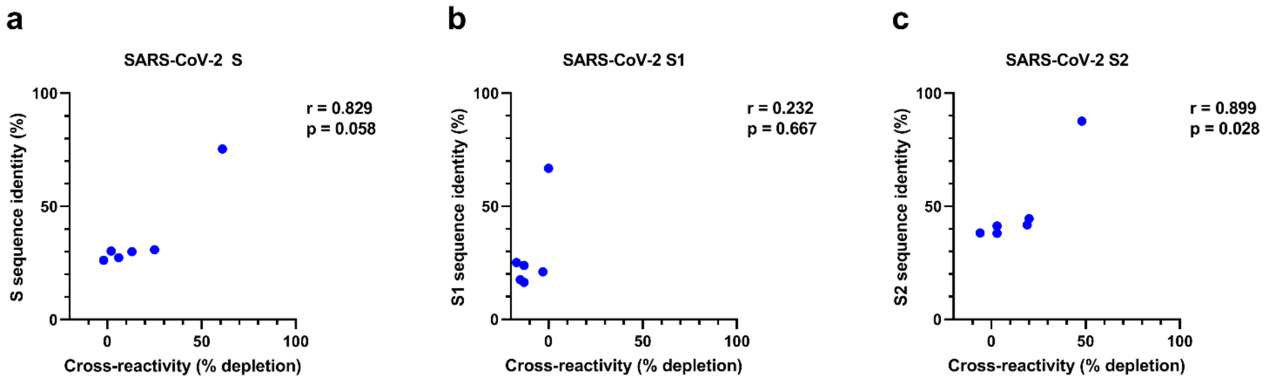


**Supplementary Figure 4. Correlation between sequence identity and reduction of cross-reactive antibodies in depletion assay for S, S1 and S2**

The correlation between S protein (a), S1 subdomain (b) and S2 subdomain (c) sequence identity of SARS-CoV-2 with the different hCoV S proteins (also shown in Supplementary Table 2) and the reduction of antibodies binding to hCoV S proteins in the SARS-CoV-2 S protein depletion assay (also shown in Figure 2b) was determined using Spearman correlation. S = spike protein, r = Spearman's rank correlation coefficient.

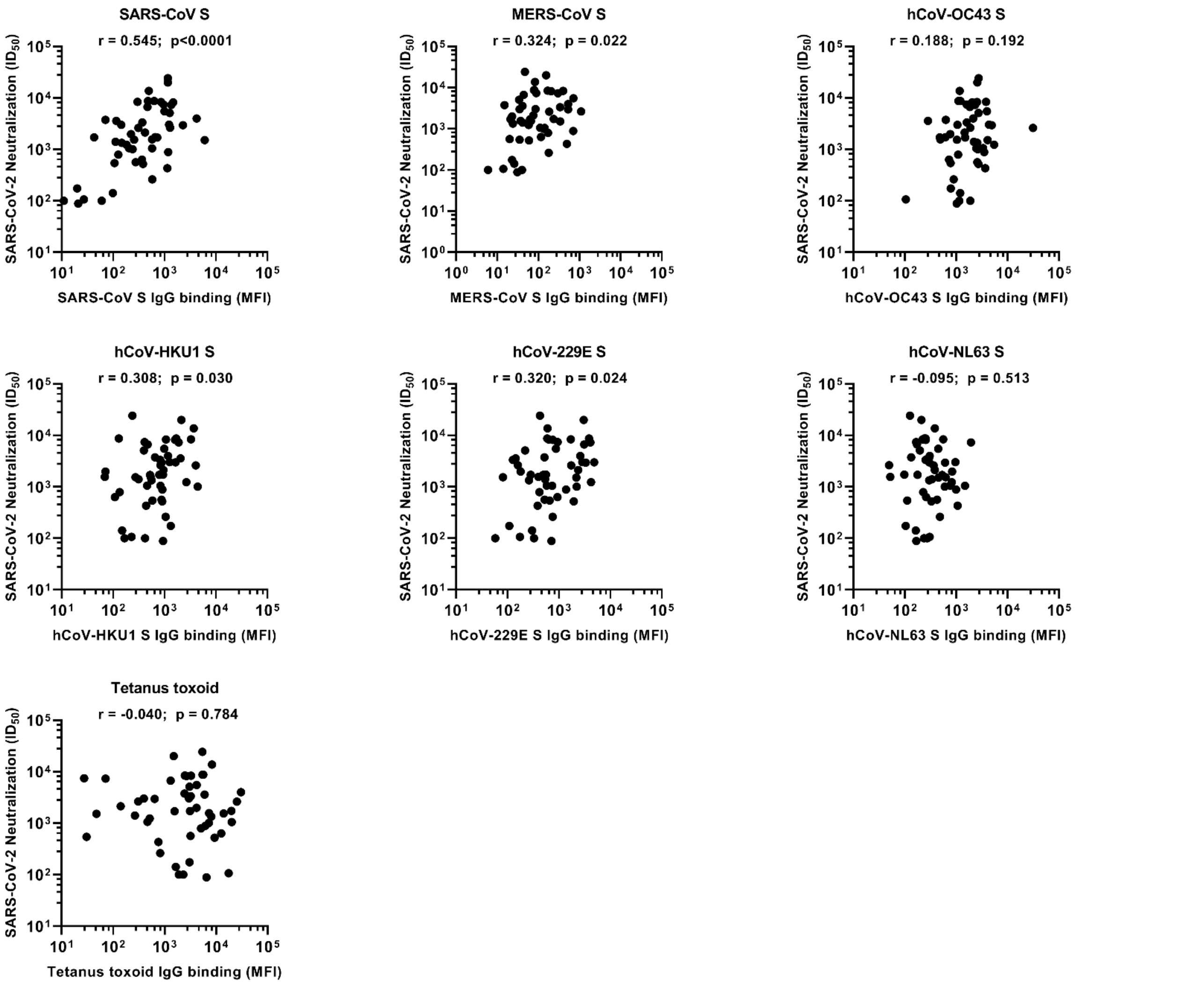


**Supplementary Figure 5. Spearman correlations between SARS-CoV-2 neutralization and antibody reactivity to hCoV S and Tetanus toxoid.**

The correlation between the SARS-CoV-2 pseudovirus neutralization (also shown in Fig. 1b) and binding IgG binding measured with a custom Luminex assay (also shown in Fig. 2a) in sera of convalescent COVID-19 patients (n = 50) was determined using Spearman correlation. MFI = median fluorescent intensity, S = spike protein, $ID_{50}$ = serum dilution at which 50% of pseudovirus is neutralized, r = Spearman's rank correlation coefficient.

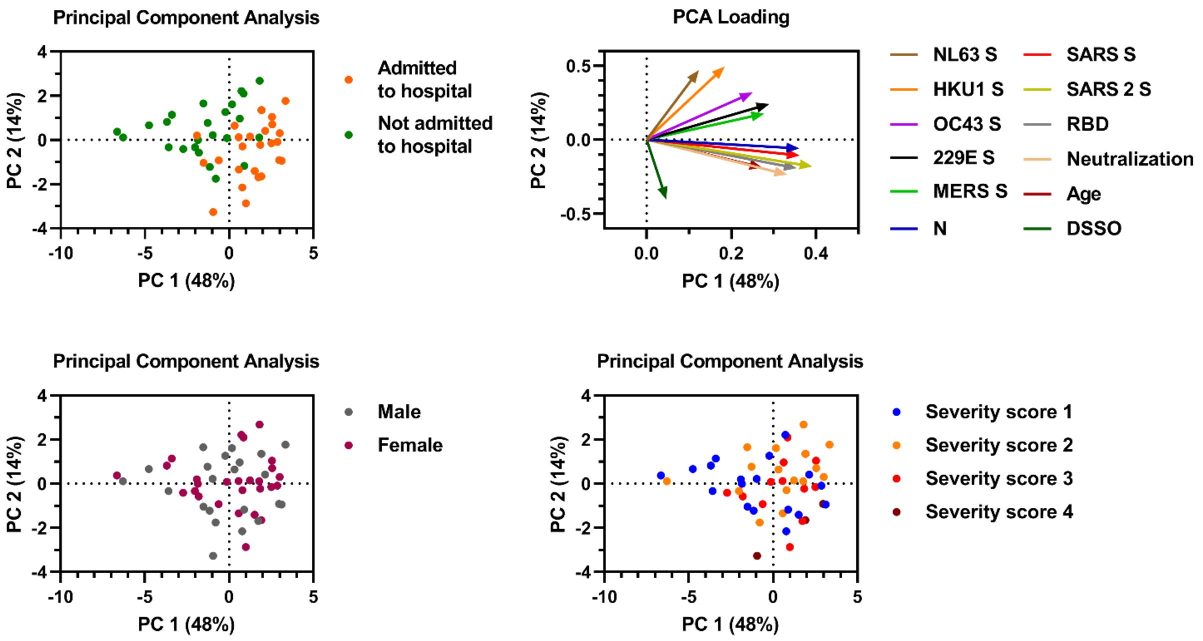


**Supplementary Figure 6. Principal Component Analysis including SARS-CoV-2 binding antibodies, neutralization, clinical characteristics and cross-reactivity.**

The Principal Component Analysis was performed using the following variables: IgG binding to all hCoV S proteins including SARS-CoV-2, IgG binding to SARS-CoV-2 RBD and nucleocapsid protein (N), SARS-CoV-2 neutralization, patient age and days since symptom onset (DSSO). Color indicates the difference between patients that needed hospitalization (orange) or stayed in home quarantine (green) (top left), between males (grey) and females (purple) (bottom left) and between patients with different WHO severity scores (bottom right, description of scoring is found in Supplementary Table 1). The loading of each variable for Principal Component (PC) 1 and 2 are shown (top right).

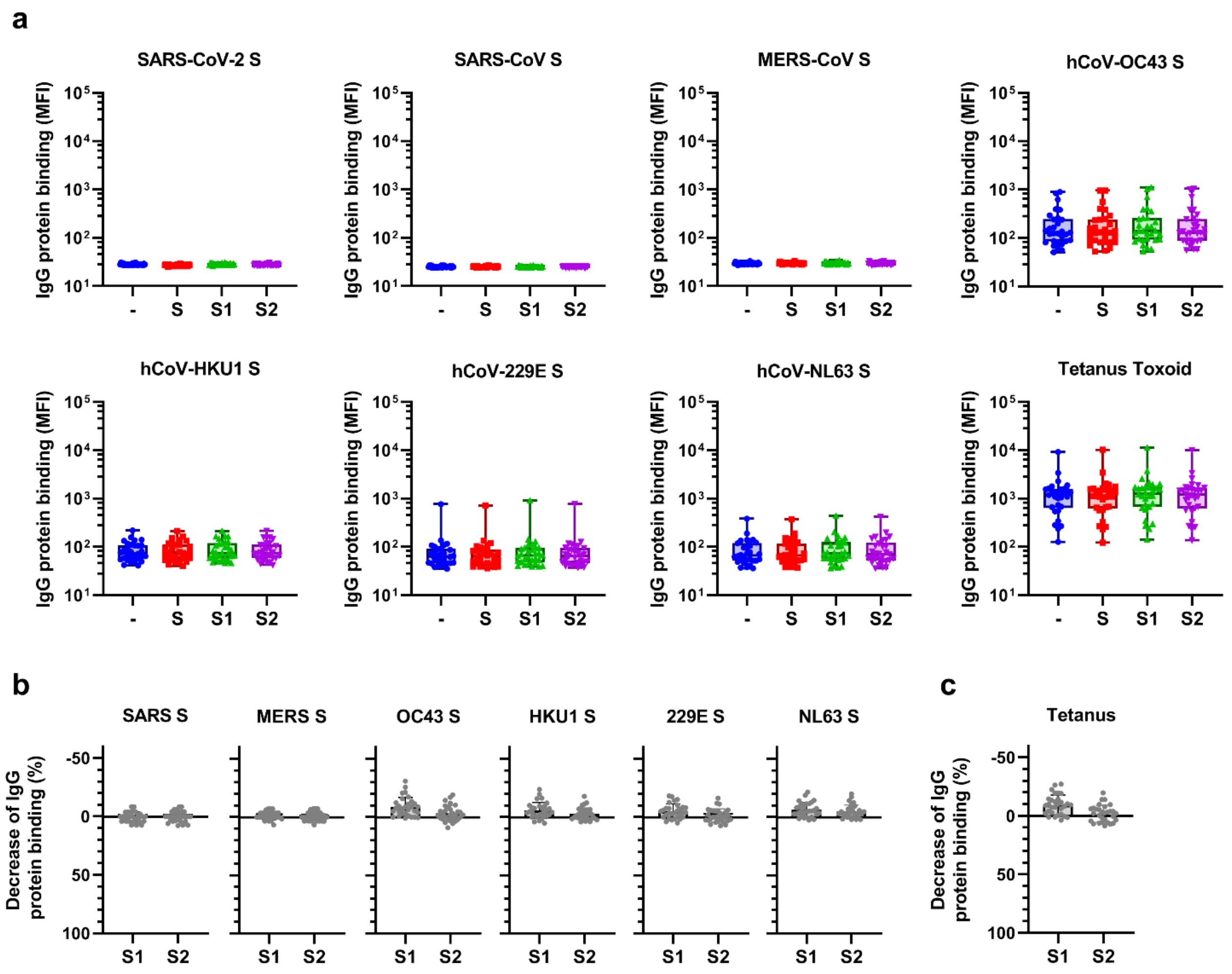


**Supplementary Figure 7. Raw data and control data from depletion assays on healthy donor sera.**

(A) MFI values of IgG binding to all hCoV S proteins (top and middle row) and control proteins (bottom row) measured with a custom Luminex assay in sera of pre-pandemic healthy donors (n = 30) after depletion with soluble recombinant SARS-CoV-2 S protein, S1 subdomain and S2 subdomain. Box plots range the minimum and maximum values. - = undepleted sera, S = trimeric S protein depleted sera, S1 = S1 subdomain depleted sera, S2 = S2 subdomain depleted sera. (B) Percent decrease of IgG binding to hCoV S proteins in sera of healthy donors (n = 30) after depletion with soluble recombinant SARS-CoV-2 S1 subdomain or S2 subdomain. (C) Percent decrease of IgG binding to Tetanus toxoid control protein in sera of healthy donors (n = 30) after depletion with soluble recombinant SARS-CoV-2 S1 subdomain or S2 subdomain. Bars represent the mean decrease of binding antibodies as percentage of the total binding IgG observed in undepleted sera of all healthy donors and error bars represent the standard deviation. Dots represent the percent decrease of binding IgG found in individual sera. MFI = median fluorescent intensity, S = spike protein.

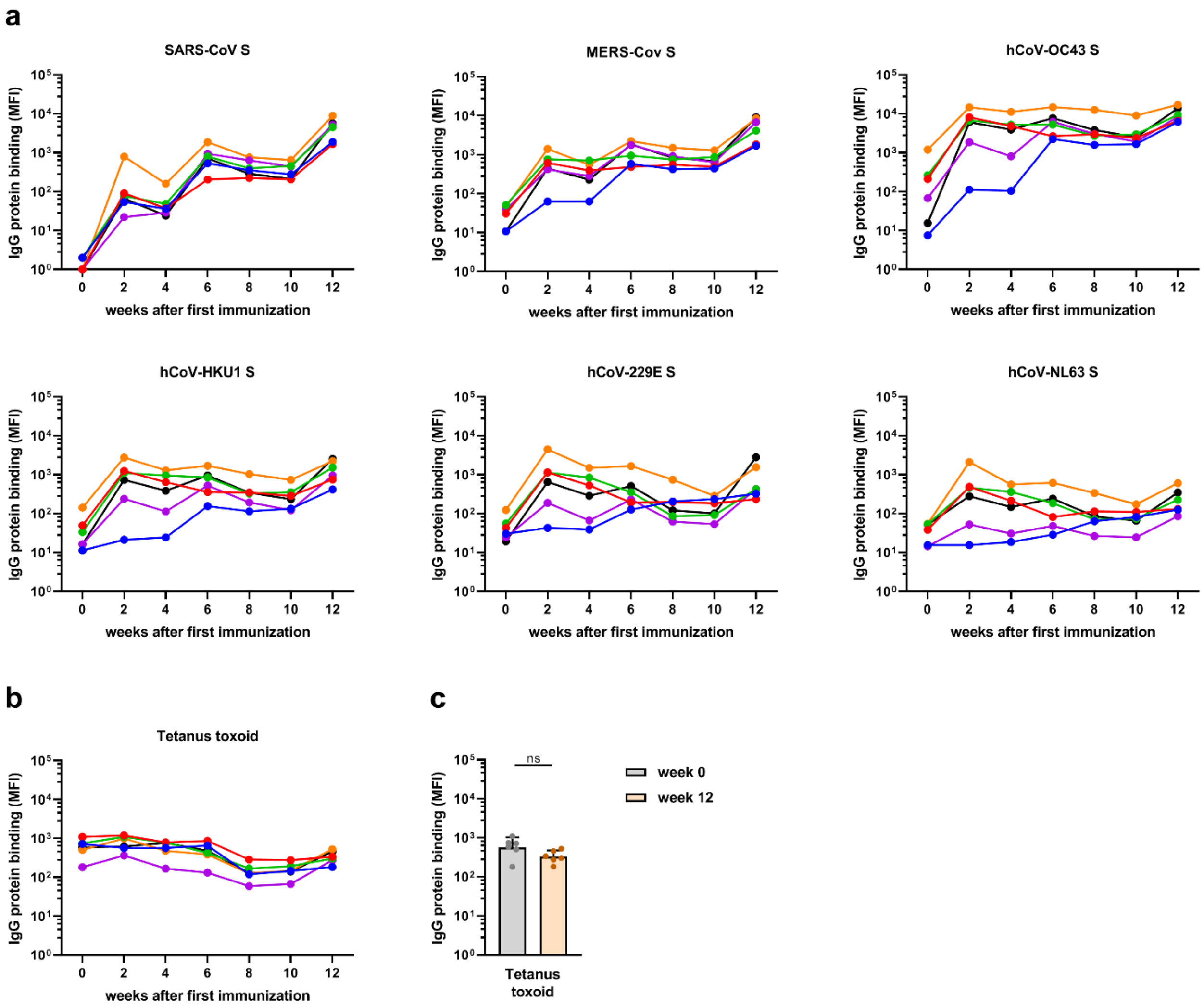


**Supplementary Figure 8. Antibodies binding to hCoV S and tetanus toxoid in immunized cynomolgus macaques over time.**

(A) IgG response over time in serum of six cynomolgus macaques immunized at week 0, 4 and 10 with a SARS-CoV-2 Spike nanoparticle vaccine, measured at a 1:50,000 serum dilution for SARS-CoV-2 and SARS-CoV S and at 1:500 serum dilution for MERS-CoV, hCoV-OC43, hCoV-HKU1, hCoV-229E and hCoV-NL63-CoV S. (B) IgG binding to Tetanus toxoid control protein, measured in 1:500 diluted serum. All MFI values are background corrected by subtraction of beads and buffer only wells. (C) IgG binding to Tetanus toxoid control protein at week 0 (pre-immunization baseline) and week 12 (after a total of three immunizations), presented in a bar chart for comparison with Fig. 4a. ns = not significant, MFI = median fluorescent intensity, S = spike protein.

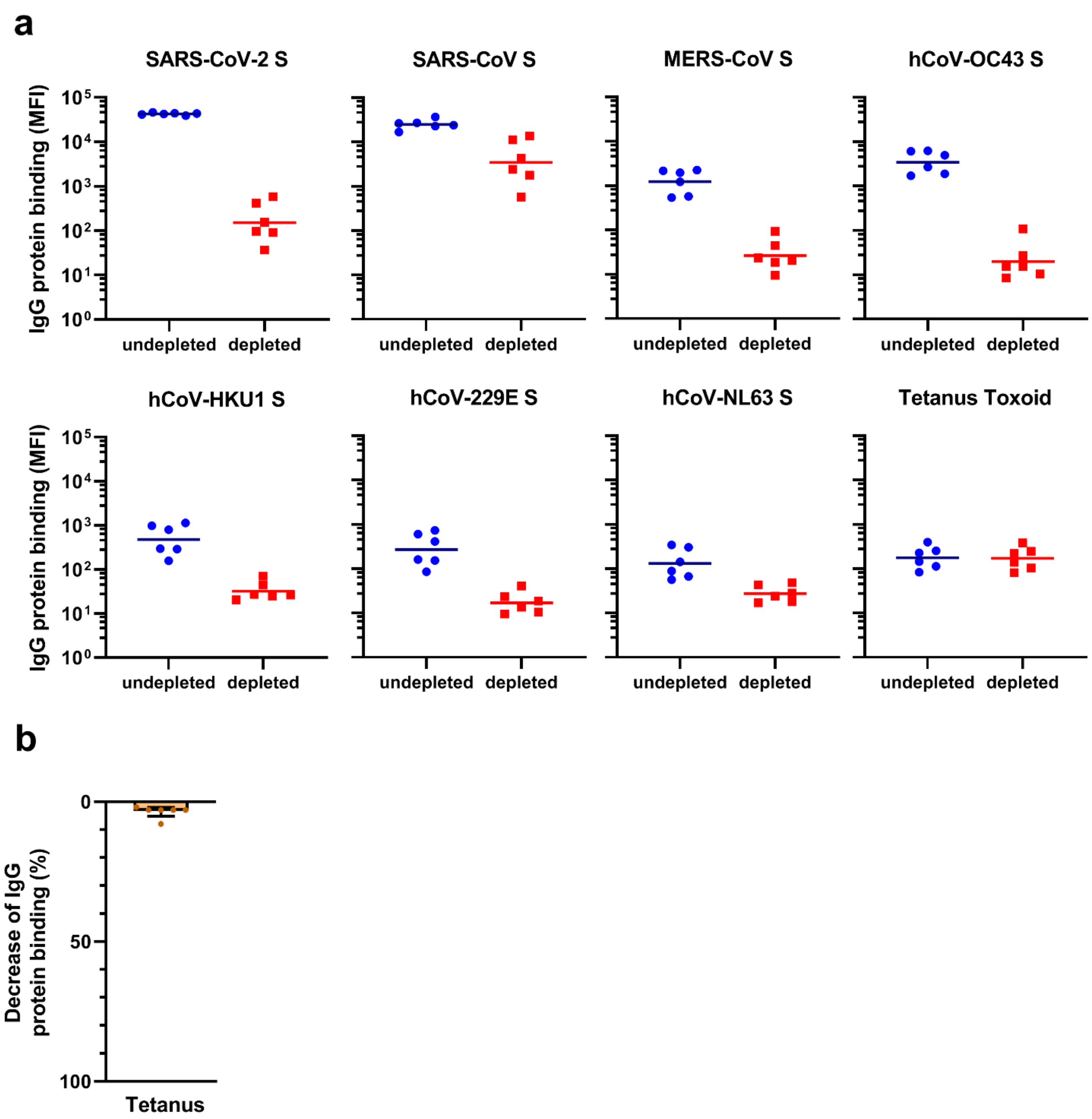


**Supplementary Figure 9. Reactivity and depletion of antibodies to hCoV S proteins and Tetanus Toxoid in immunized cynomolgus macaques.**

(A) MFI values of IgG binding to all hCoV S proteins and Tetanus toxoid control protein with and without depletion with soluble recombinant SARS-CoV-2 S protein, measured by Luminex assay in 1:50,000 diluted serum (for SARS-CoV-2 and SARS-CoV) or 1:500 diluted serum (for all other proteins) for six cynomolgus macaques immunized at week 0, 4 and 10 with a SARS-CoV-2 S protein nanoparticle vaccine. The line indicates the geometric mean. (B) Percent decrease of IgG binding to Tetanus toxoid control protein in 1:500 diluted sera at week 12 after depletion with soluble recombinant SARS-CoV-2 S protein, presented in bar charts for comparison with Fig. 4b. Bars represent the mean decrease of binding IgG as percentage of the total binding IgG observed in undepleted sera of the six cynomolgus macaques. Dots represent the percent decrease of binding IgG in each individual animal. MFI = median fluorescent intensity, S = spike protein.

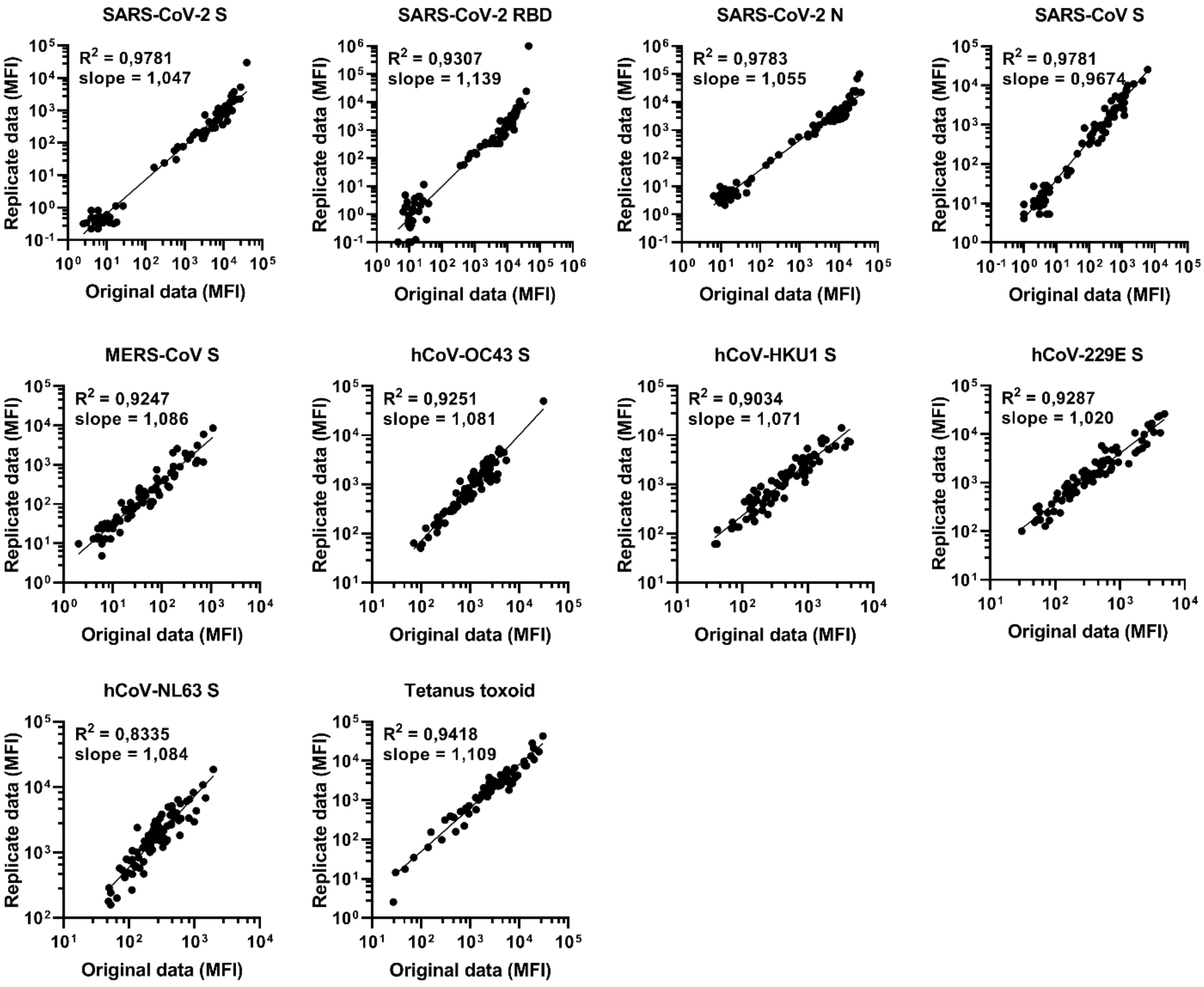


**Supplementary Figure 10. Reproducibility of the Luminex assay.**

The reproducibility of the custom Luminex assay is shown by plotting data from two independent assays performed on different days with the same samples. The data on the Y axis is found in Fig. 3 and the data on the X axis is also found in Fig. 1 and 2. The quality of the replicates is presented by the $R^2$ and slope of a simple linear regression performed on the $log_{10}$ transformed MFI data. MFI = median fluorescent intensity, S = spike protein, N = nucleocapsid, RBD = receptor binding domain.
